# Implementation of SARS-CoV-2 monoclonal antibody infusion sites at three medical centers in the United States: Strengths and challenges assessment to inform COVID-19 pandemic and future public health emergency use

**DOI:** 10.1101/2021.04.05.21254707

**Authors:** Anastasia S. Lambrou, John T. Redd, Miles A. Stewart, Kaitlin Rainwater-Lovett, Jonathan K. Thornhill, Lynn Hayes, Gina Smith, George M. Thorp, Christian Tomaszewski, Adolphe Edward, Natalia Elías Calles, Mark Amox, Steven Merta, Tiffany Pfundt, Victoria Callahan, Adam Tewell, Helga Scharf-Bell, Samuel Imbriale, Jeffrey D. Freeman, Michael Anderson, Robert P. Kadlec

## Abstract

**Background:** The COVID-19 pandemic caught the globe unprepared without targeted medical countermeasures, such as therapeutics, to target the emerging SARS-CoV-2 virus. However, in recent months multiple monoclonal antibody therapeutics to treat COVID-19 have been authorized by the U.S. Food and Drug Administration (FDA) under Emergency Use Authorization (EUA). Despite these authorizations and promising clinical trial efficacy results, monoclonal antibody therapies are currently underutilized as a treatment for COVID-19 across the U.S. Many barriers exist when deploying a new infused therapeutic during an ongoing pandemic with limited resources and staffing, and it is critical to better understand the process and site requirements of incorporating monoclonal antibody infusions into pandemic response activities.

**Methods:** We examined the monoclonal antibody infusion site process components, resources, and requirements during the COVID-19 pandemic using data from three initial infusion sites at medical centers in the U.S. supported by the National Disaster Medical System. A descriptive analysis was conducted using process assessment metrics to inform recommendations to strengthen monoclonal antibody infusion site implementation.

**Results:** The monoclonal antibody infusion sites varied in physical environment and staffing models due to state polices, infection control mechanisms, and underlying medical system structure, but exhibited a common process workflow. Sites operationalized an infusion process staffing model with at least two nurses per ten infusion patients. Monoclonal antibody implementation success factors included tailoring the infusion process to the patient community, strong engagement with local medical providers, batch preparing the therapy before patient arrival, placing the infusion center in proximity to emergency services, and creating procedures resilient to EUA changes. Infusion process challenges stemmed from confirming patient SARS-CoV-2 positivity, strained staff, scheduling needs, and coordination with the pharmacy for therapy preparation.

**Conclusions:** Infusion site processes are most effective when integrated into the pre-existing pandemic response ecosystems and can be implemented with limited staff and physical resources. As the pandemic and policy tools such as EUAs evolve, monoclonal antibody infusion processes must also remain adaptable, as practice changes directly affect resources, staffing, timing, and workflows. Future use may be aided by incorporating innovative emergency deployment techniques, such as vehicle and home-based therapy administration, and by developing drug delivery mechanisms that alleviate the need for observed intravenous infusions by medically-accredited staff.

## BACKGROUND

Severe acute respiratory syndrome coronavirus (SARS-CoV-2) emerged in late 2019 and ignited a global pandemic with detrimental impacts on health systems across the world. This novel virus caught the globe unprepared without targeted medical countermeasures (MCMs), such as therapeutics, to treat individuals with coronavirus 2019 (COVID-19). As the pandemic progressed and scientific progress was rapidly stimulated, the therapeutic toolkit to treat COVID-19 evolved to include monoclonal antibodies.^1^ Monoclonal antibody therapeutics to treat COVID-19 are composed of laboratory-synthesized SARS-CoV-2 neutralizing antibodies, most often isolates from infected individuals, isolated for specific immunologic properties such as binding, neutralization, and effector functions.^2^ Since November 2020, multiple formulations of monoclonal antibodies have been authorized by U.S. Food and Drug Administration’s (FDA) under Emergency Use Authorization (EUA).^3^ Recent clinical trials on monoclonal antibody therapies suggest that early use of these drugs can reduce COVID-19 symptom severity, SARS-CoV-2 viral load, and hospitalization in infused outpatient populations as compared to individuals given placebos.^4–6^ Real-world effectiveness studies have also provided evidence that monoclonal antibody infusions reduce hospitalization rates in high risk patient populations.^7-9^ These monoclonal antibody therapies are currently administered as intravenous infusions to treat individuals with mild to moderate COVID-19. The EUAs also specify monoclonal antibody infusion eligibility requirements for potential patients at high risk for COVID-19 complications, such as age, BMI, and pre-existing conditions (SI Table 1). EUAs are regulatory tools used during public health emergencies, such as pandemics, to expand use, system implementation, and further study of new therapeutics.^10^

Despite the EUAs and promising clinical trial results, monoclonal antibody therapies are currently underutilized as a treatment for COVID-19 across the U.S. This is hypothesized to be due to gaps in outreach to both providers and patient communities, strict EUA criteria, and infusion site implementation barriers during the ongoing pandemic, such as staffing, resources, and infection control.^11^ Incorporating monoclonal antibodies into COVID-19 response efforts may relieve stress on medical centers through reducing disease severity and hospitalizations.^12^ Monoclonal antibody use is increasing in some settings across the U.S., but there is limited research on the implementation of this therapy, resources needed to maintain an infusion site, and lessons learned to inform the scale-up of this pandemic response tool. Monoclonal antibody therapeutics may also play a critical role in future emerging biological threats, including the newly-described emerging variant SARS-CoV-2 isolates, as they can be rapidly manufactured and can be used as a treatment before other MCMs, such as vaccines, are evaluated and distributed.^13^ Vaccines may also require multiple weeks or doses to elicit protection, while monoclonal antibodies serve as a treatment to reduce the burden of a novel pathogen. It is critical to learn from the ongoing implementation of monoclonal antibody infusions during the COVID-19 pandemic to inform the scale-up of this therapy, and other biologics, during the current and future emergencies.

The purpose of this investigation was to describe monoclonal antibody infusion site implementation and requirements during the COVID-19 pandemic using data from three sites in the U.S. supported by the Office of the Assistant Secretary for Preparedness and Response (ASPR). A set of standard metrics was utilized to evaluate site infusion process staffing model, resources, strengths and challenges. Diagrams of the monoclonal antibody infusion process components and infusion site physical environment illustrate various therapy implementation layouts. The descriptive metrics analysis informs the implementation of a monoclonal antibody infusion site for the COVID-19 pandemic response efforts and for future use to tackle emerging infectious disease threats. This is a critical window during the pandemic in the U.S. to examine the implementation of monoclonal antibody infusion sites for outpatients as the response is currently marked by recent therapy EUAs and the steadily growing mass distribution of COVID-19 vaccines.

## METHODS

### Infusion Sites

Data were collected from three medical centers in the United States (U.S.), El Centro Regional Medical Center (El Centro, CA), TMC HealthCare (Tucson, AZ), and Sunrise Hospital and Medical Center (Las Vegas, NV) between January and February 2021. These sites recently implemented monoclonal antibody infusions during the pandemic to treat individuals with mild and moderate COVID-19 using EUA criteria and by collaborating with ASPR’s National Disaster Medical System (NDMS) Disaster Medical Assistance Teams (DMATs). All three medical sites then transitioned to maintaining their own monoclonal antibody infusion sites without ASPR support and incorporated monoclonal antibody infusion into their COVID-19 pandemic response workflows. This investigation was concerned with describing the infusion site process workflows after the DMAT teams departed and the medical systems transitioned their processes to ensure sustainability during the COVID-19 pandemic. These sites were selected due to their early adoption of monoclonal antibody delivery and experience in site implementation and maintenance. The three sites also exhibited diverse and underserved patient populations, process approaches, infrastructures, and physical locations to inform monoclonal antibody infusion process scale-up across the U.S. This clinical support activity was conducted as part of the ASPR public health response to the COVID-19 pandemic and at the request of the host institutions. Under HHS Office of Health Research Protection guidelines, it was judged a non-research COVID-19 response activity. The Johns Hopkins University Applied Physics Laboratory (JHU/APL) Environmental Health Services Board and all three medical sites also deemed this work non-human subjects research exempt from institution review board approval.

### Data Collection

Data were collected through three mechanisms to inform the monoclonal antibody infusion process assessment, model, and recommendations: 1) key informant interviews, 2) onsite observations, and 3) infusion records. A process assessment framework informed the seven key metrics on which data were collected to ensure standard data collection at each site (Figure 1): logistics, timing, staffing, physical environment, resources, monitoring and resilience, and engagement (SI Table 2). The seven framework metrics describe critical quantitative and qualitative characteristics of the infusion process to inform the assessment and propose future recommendations.

**Figure 1.**
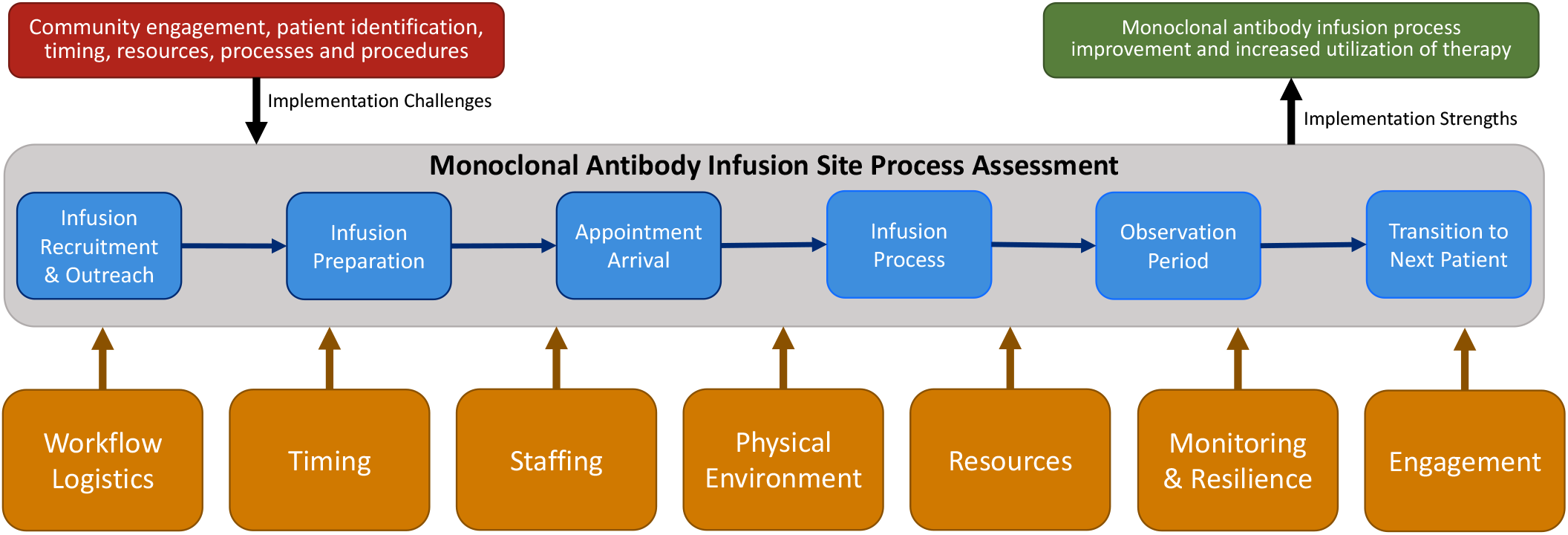
Monoclonal antibody infusion site process assessment framework and metrics to examine the strengths and challenges related to implementation.

Semi-structured key informant interviews were conducted at each site using an interview guide to collect data on infusion process assessment metrics to ensure standard data collection. Interviews were conducted with the medical center’s Chief Medical Officer (CMO), infusion site logistics lead, infection control lead, director of pharmacy, and infusion site staff. Each of the three different medical centers’ monoclonal antibody infusion sites were visited by the study team to observe and map the infusion process workflow. Each step in the infusion process was timed for multiple patients and the staff, resources, and information needed for the step were recorded. The onsite observations also facilitated validating data from the key informant interviews.

### Descriptive Analyses

Descriptive analysis of the monoclonal antibody infusion process was conducted to examine the timing, staffing needs, resources, and information flow of each component of the process. The process was examined from patient engagement through the infusion appointment and discharge from the infusion site. The physical environment of each infusion site was also mapped to analyze resource and implementation needs for this new therapy option. Data on each process metric from the process assessment framework was synthesized and compiled for each site.

## RESULTS

### Infusion Site Process Workflow

A descriptive analysis of three medical center monoclonal antibody infusion sites was conducted using a process assessment to inform recommendations to strengthen infusion site implementation during current pandemic response efforts. This investigation evaluated the process of monoclonal antibody infusion and staffing equipment, physical space, and resource requirements during the COVID-19 pandemic. A general monoclonal antibody infusion site workflow process (Figure 2) was developed to integrate the data from the three data collection sites. It is important to note that there was not a single standard monoclonal antibody infusion site process workflow. Each site exhibited common process components, staffing models, and resources, yet adapted the system to address local policies, patient populations, and medical center characteristics. An effective monoclonal antibody infusion site optimized the volume of infused patients and minimized patient appointment time and stress on the underlying medical system.

**Figure 2.**
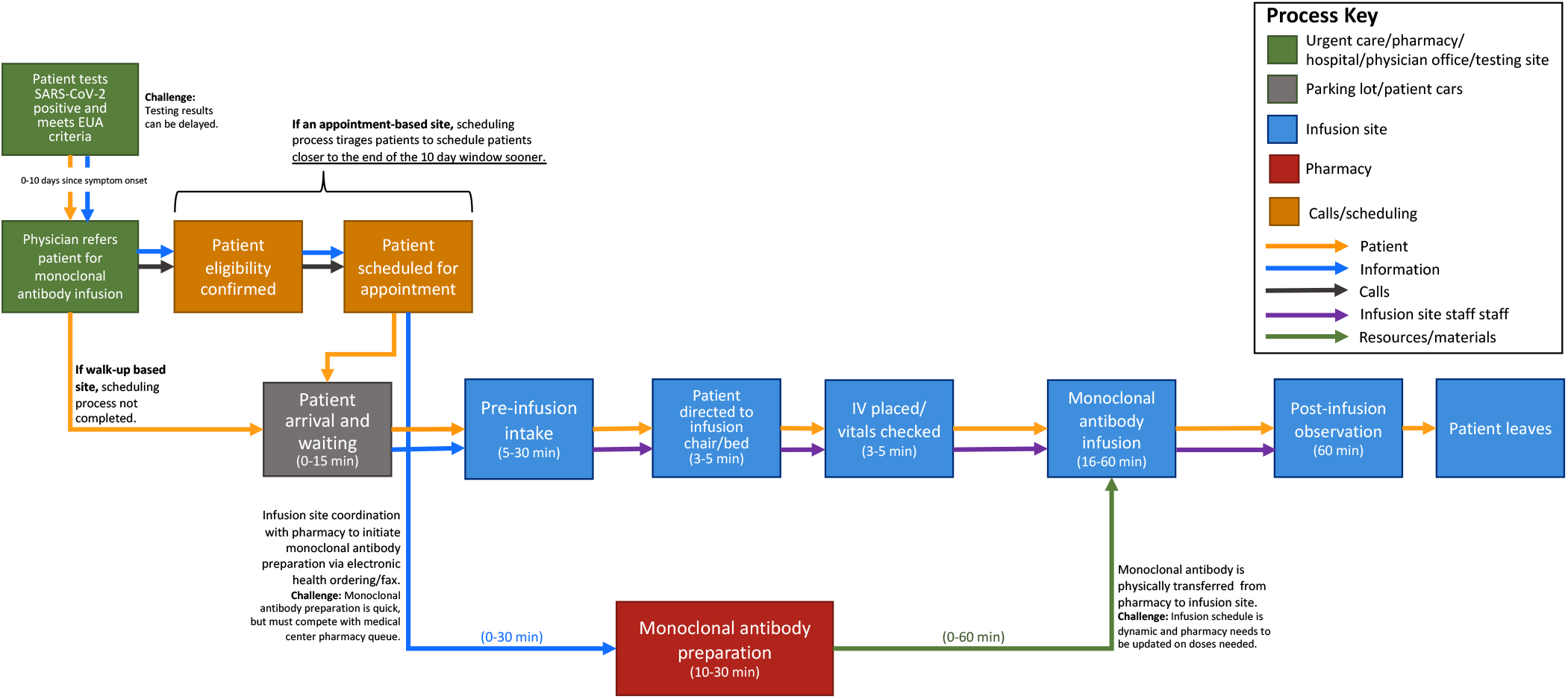
General monoclonal antibody infusion site process workflow examining the network of physical environments, patients, information, calls, staff, and resources, informed by the workflows and assessments of each data collection site.

**Figure 3.**
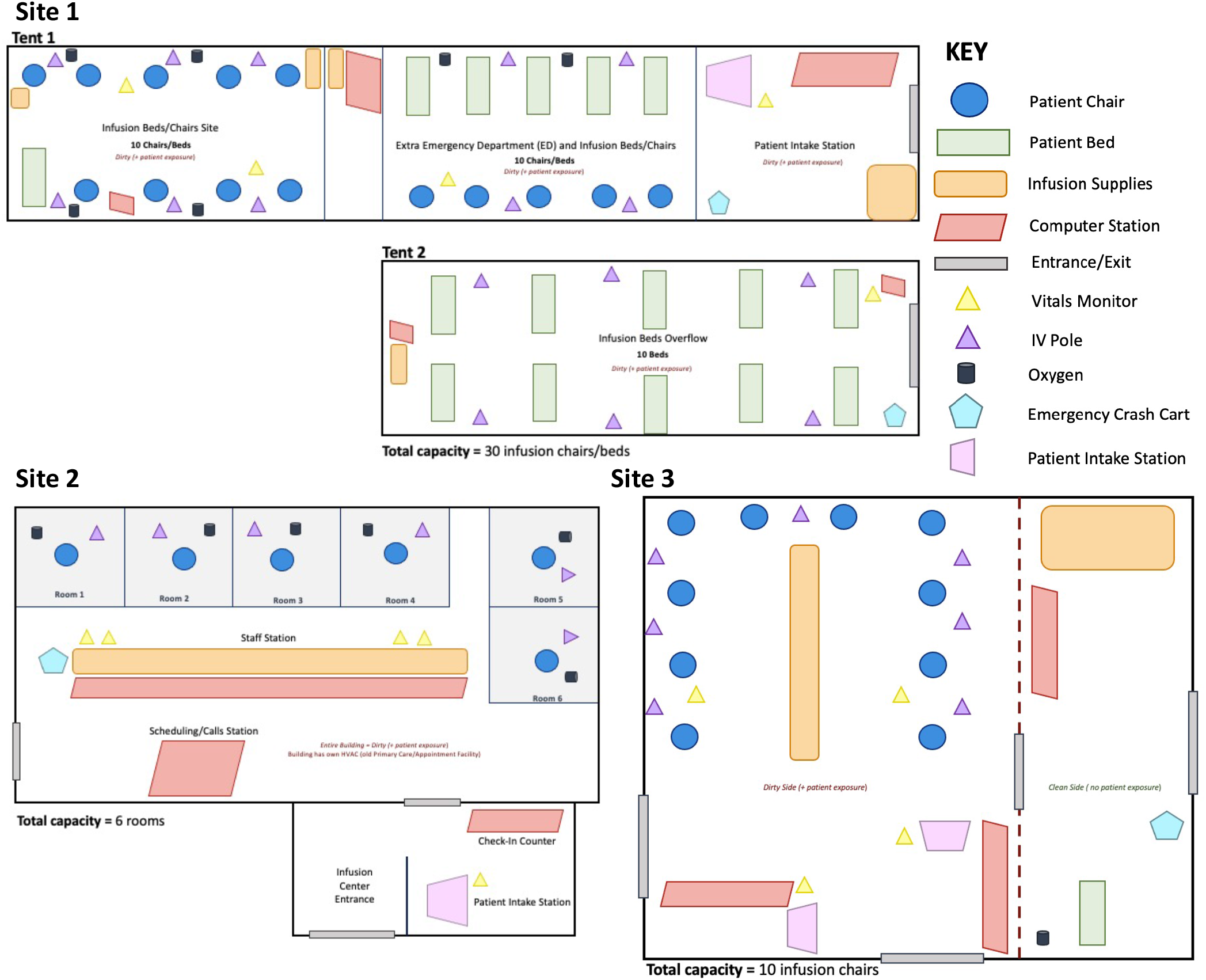
Monoclonal antibody infusion site physical environment schematics of Sites 1-3 indicating resources, site type, and layout.

The sites exhibited two major medical center mechanisms of implementing a monoclonal antibody infusion site: 1) an outpatient infusion clinic model, and 2) an Emergency Department (ED) medication visit model (Table 1). Site 1 employed a model tied to ED operations, while Sites 2 and 3 operated as outpatient infusion sites co-located with a medical center. The infusion sites also presented two appointment types: 24/7 walk-up and scheduled appointments during business hours. The three sites started infusions at different times: first Site 1 started on November 17th, 2020, and Sites 2 and 3 initiated infusions the same week, respectively on January 7^th^ and 8^th^, 2021. Site 1 completed 636 infusion since starting the site with an average rate of 6 infusions per day. Site 2 recorded the highest number of infusions with 824 patients infused, amounting to a rate of approximately 16 infusions per day. Lastly, Site 3 completed 402 infusions with a rate of 8 patients infused per day.

**Table 1.** Monoclonal antibody infusion process logistics and timing metrics from the three National Disaster Medical System-supported infusion sites and related strengths and challenges to inform implementation.

| Logistics and Timing Metrics | Site 1 | Site 2 | Site 3 | Implementation Considerations |  |
| --- | --- | --- | --- | --- | --- |
|  |  |  |  | Strengths | Challenges |
| Infusion Site Type | Walk-up tent infusion site | Appointment-based outpatient infusion site | Appointment-based tent infusion site | Walk-up sites were beneficial in communities with low healthcare system engagement<br>Appointment-based sites facilitated batch preparation of monoclonal antibody infusion doses, shortening the overall time of the appointment<br>30-minute staggering between patient group arrivals improved patient flow due to 15-30 minute intake process | Walk-up sites exhibited longer wait times for on-demand pharmacy preparation of the monoclonal antibody<br>Batch preparation of monoclonal antibodies resulted in unused doses for walk-up systems<br>Walk-up site had large variability in timing due to confirming the patient's SARS-CoV-2 positivity upon arrival<br>Appointment-based sites required increased staffing and planning to schedule patients |
| Process Type | Emergency medical visit | Outpatient infusion procedure | Outpatient infusion procedure |  |  |
| Infusion Site Start Date | Nov 17, 2020 | Jan 7, 2021 | Jan 8, 2021 |  |  |
| Total Patients Infused during Study Period<br>(Start-Feb 26 2021) | 636 | 824 | 402 |  |  |
| Average Rate (Patients/Day) | 6 | 16 | 8 |  |  |
| Most Significant Logistics Barriers | Confirming SARS-CoV-2 patient positivity criteria<br>Coordination with pharmacy for monoclonal antibody preparation | Coordination with pharmacy for monoclonal antibody preparation<br>Staffing needs for scheduling process | Coordination with pharmacy for monoclonal antibody preparation<br>Staffing needs for scheduling process |  |  |
| Hours of Operation | 24 hours/day<br>• 7 days a week | Monday-Friday<br>• 9:00am-5:00pm | Monday-Friday<br>• 9:00am-5:00pm |  |  |

Generally, the process components were initiated by a prospective patient testing positive for SARS-CoV-2, and with scheduling-based infusion sites, patients having first to obtain a provider referral for monoclonal antibody treatment with confirmation that they meet the EUA criteria Robust and timely local SARS-CoV-2 test result turnaround was critical to effective monoclonal antibody implementation, as the current EUA requires the infusion to occur within 10 days of symptom onset in patients with a documented positive COVID viral test result. Areas with SARS-CoV-2 testing turnaround close to one week delayed patient referral and created monoclonal antibody uptake obstacles. Infusion site appointments had three major components. The first component was a pre-infusion intake process to confirm patient eligibility, collect vitals, obtain patient consent, and insert an IV. The next component was the monoclonal antibody infusion process, which ranged from 16-60 minutes depending upon the specific therapy available and size of infusion bags. This time was EUA-dependent and this process must remain flexible to changes in infusion requirements, as the guidelines changed from 60 to 16 minutes during the study period. The last component was the EUA-specified 60-minute patient observation period of each patient to monitor for any adverse events.

Three process components contributed the most to patient visit time variability: 1) scheduling appointments, 2) pre-infusion patient intake, and 3) monoclonal antibody coordination with the medical center pharmacy. These three process components also created stresses on already constrained staffing resources. A critical barrier of the infusion process at each of the three sites was the pharmacy’s preparation of the monoclonal antibody and coordination with the infusion site on therapy doses and timing. Scheduling-based infusion site pharmacies were equipped with data to enable pre-preparation of monoclonal antibody doses in batches before patients arrive. The three infusion sites emphasized that coordination with the pharmacy is difficult due to physical proximity and the need to conserve any prepared doses. Monoclonal antibody infusion process workflows were strongly shaped by EUA requirements regarding drug preparation, storage, timing, and delivery.

### Infusion Process Staffing Metrics

Similar to the infusion process components, the infusion site staffing metrics varied between sites. The different staffing models relied on the same underlying requirements to ensure monoclonal antibody referral, prescription, preparation, and administration (Table 2). Staffing models differed due to state policies and the different underlying staffing structures of the three medical centers. Each staffing model consisted of an advanced practice provider (APP) or physician, a nursing team, and a pharmacy team. The infusion site operations relied heavily on the nursing team and the more effective infusion process workflows separated the nursing team into two distinct task areas: patient pre-infusion intake tasks and the infusion-related tasks. The consistent recommendation from the infusion sites for the minimal staffing needs estimated two registered nurses (RNs) are needed for every 10 infusion patients. Informed by initial implementation experience, sites recommended developing a process workflow split into two staffing components with one RN completing pre-infusion and intake processes such as patient initial vitals, data collection, and consent. All sites also recommended integrating paramedics, to start IVs and monitor patients, into the staffing model to alleviate stress on constrained medical center nursing staff. One site leveraged a local medical volunteer organization to support staffing the infusion site during the ongoing pandemic to reduce stress on the medical center’s pandemic response staffing. Each of the three sites also strongly recommended initiating a multidisciplinary staffing meeting between the medical center’s leadership, pharmacy, infection control, ED, nursing, information technology, and security to coordinate the implementation process and medical center staffing allocation. These representatives were not needed for the day-to-day operations of the monoclonal antibody infusion site, but their expertise and support were for developing the initial workflow and staffing models at the three sites.

**Table 2.**
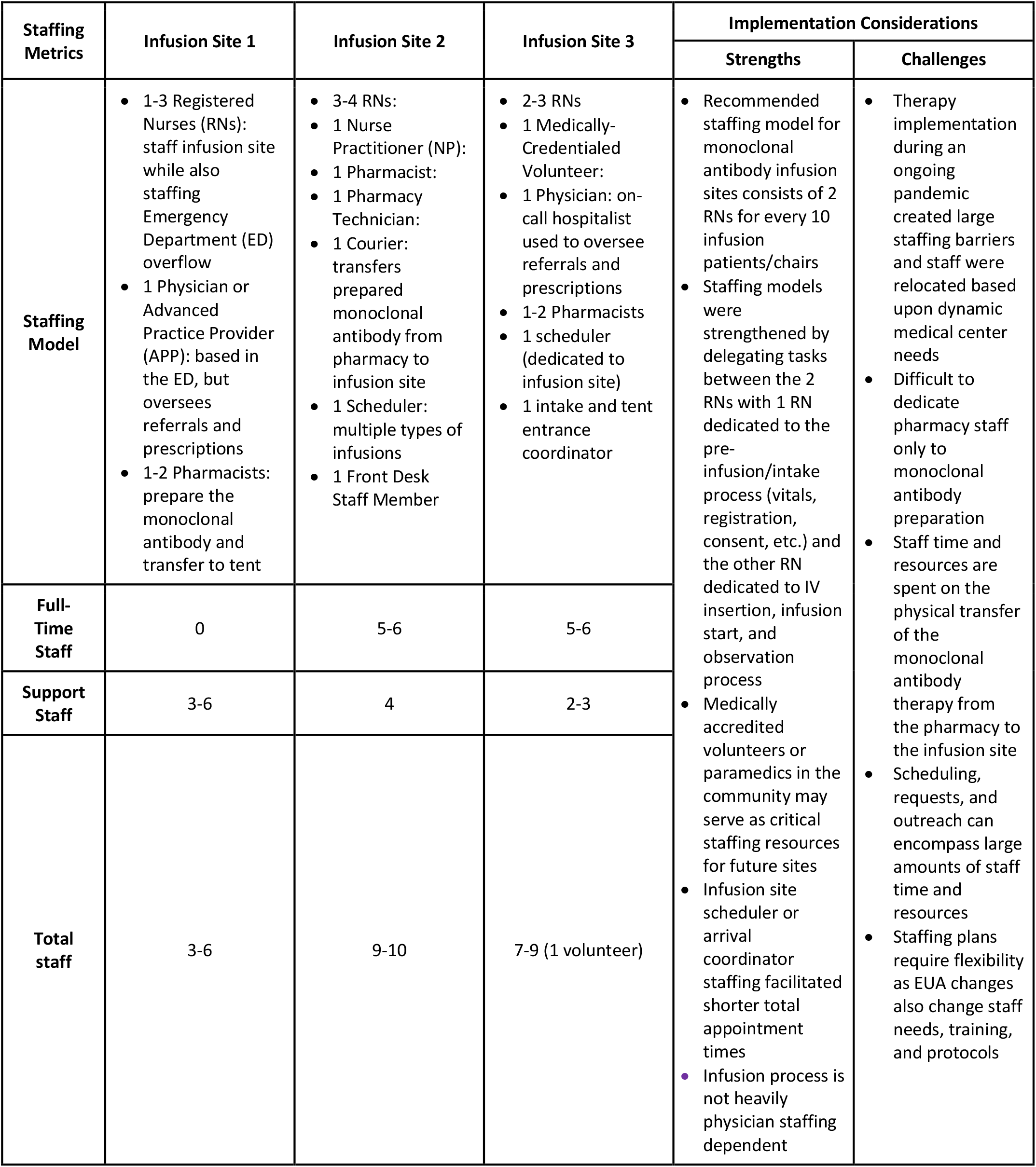
Monoclonal antibody infusion process staffing metrics from the three National Disaster Medical System-supported infusion sites and strengths and challenges related to staffing and implementation decision-making.

### Physical Environment and Resource Metrics

The different external and internal physical environments exhibited by the three monoclonal antibody infusion sites were influenced by infection control, resource transport, staffing, and emergency response plan considerations. Monoclonal antibody recipients are all laboratory-confirmed SARS-CoV-2-positive patients and likely infectious; consequently, it was critical to separate the infusion site from other medical center operations with uninfected individuals. Two of the sites created temporary tent-based infusion sites next to their ED to maintain a separate physical space and HVAC system for infection control purposes, but remain near emergency services for potential adverse events and the pharmacy for monoclonal antibody preparations. One site converted a former primary care clinic located a short distance away from the main medical center into a monoclonal antibody infusion site. This building was only being used by monoclonal antibody patients and the therapy was transferred by a driving courier from the pharmacy in the main medical center campus to the site.

The sites differed in the total number of patients who could be infused at one point in time. While the indoor site allocated six rooms for infusion, the two tent sites had 10 and 30 infusion chairs. Medical and technological infusion site resources were needed to perform the infusion process, record patient data, and ensure an infection-controlled environment. The resources did not vary greatly between the three infusion sites; however, some sites improved the overall monoclonal antibody infusion process by using a mobile, miniature refrigeration unit to store batches of the monoclonal antibody and scanners to rapidly send prescription and paperwork (Table 3). The temporary tent sites required more infrastructure resources such as electricity sources, power strips, lights, HVAC systems, and generators to remain self-sufficient while adjacent to the medical center. At the current stage in the pandemic, the three infusion sites did not report any supply chain barriers related to the physical environment and infusion-related resources.

**Table 3.**
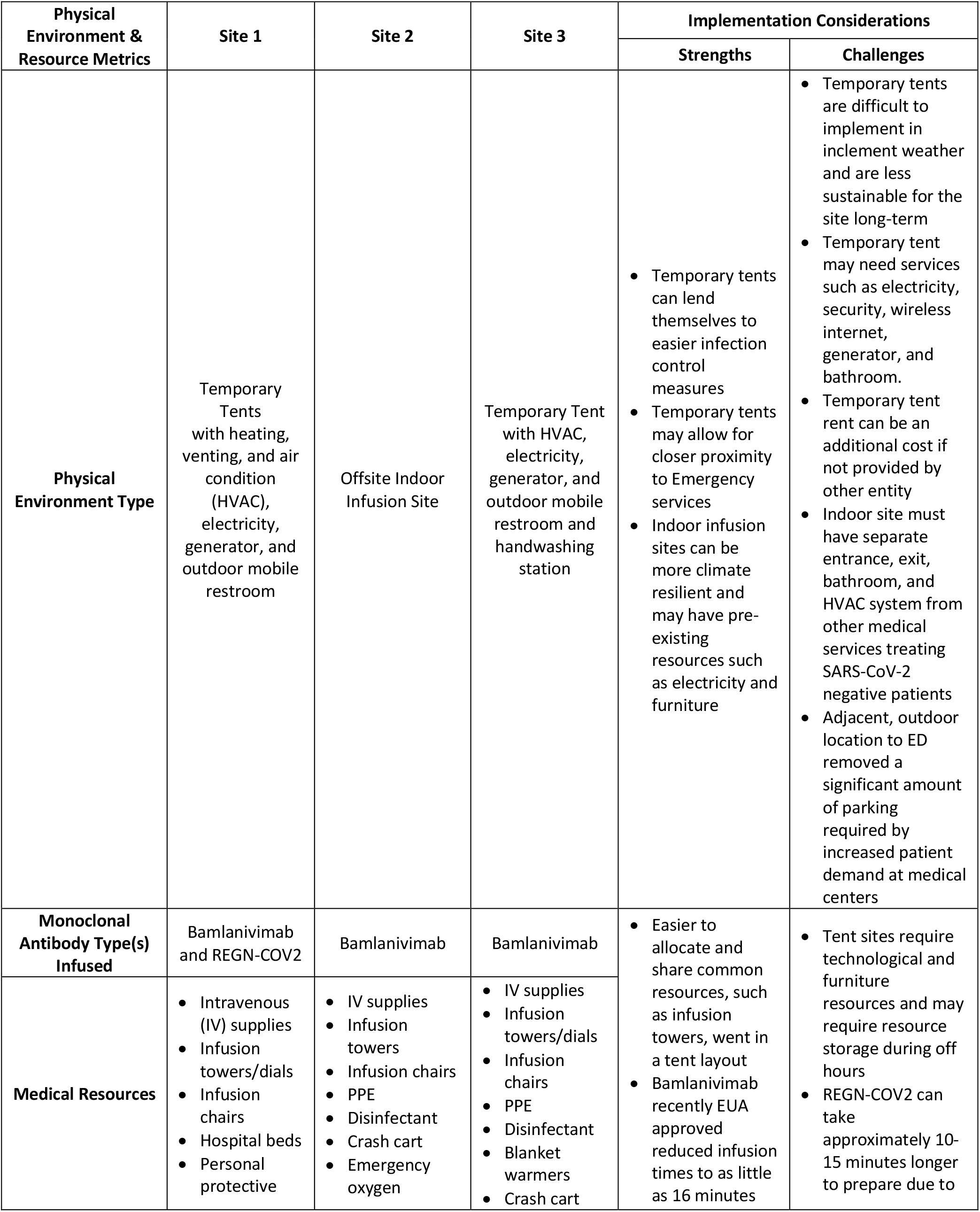
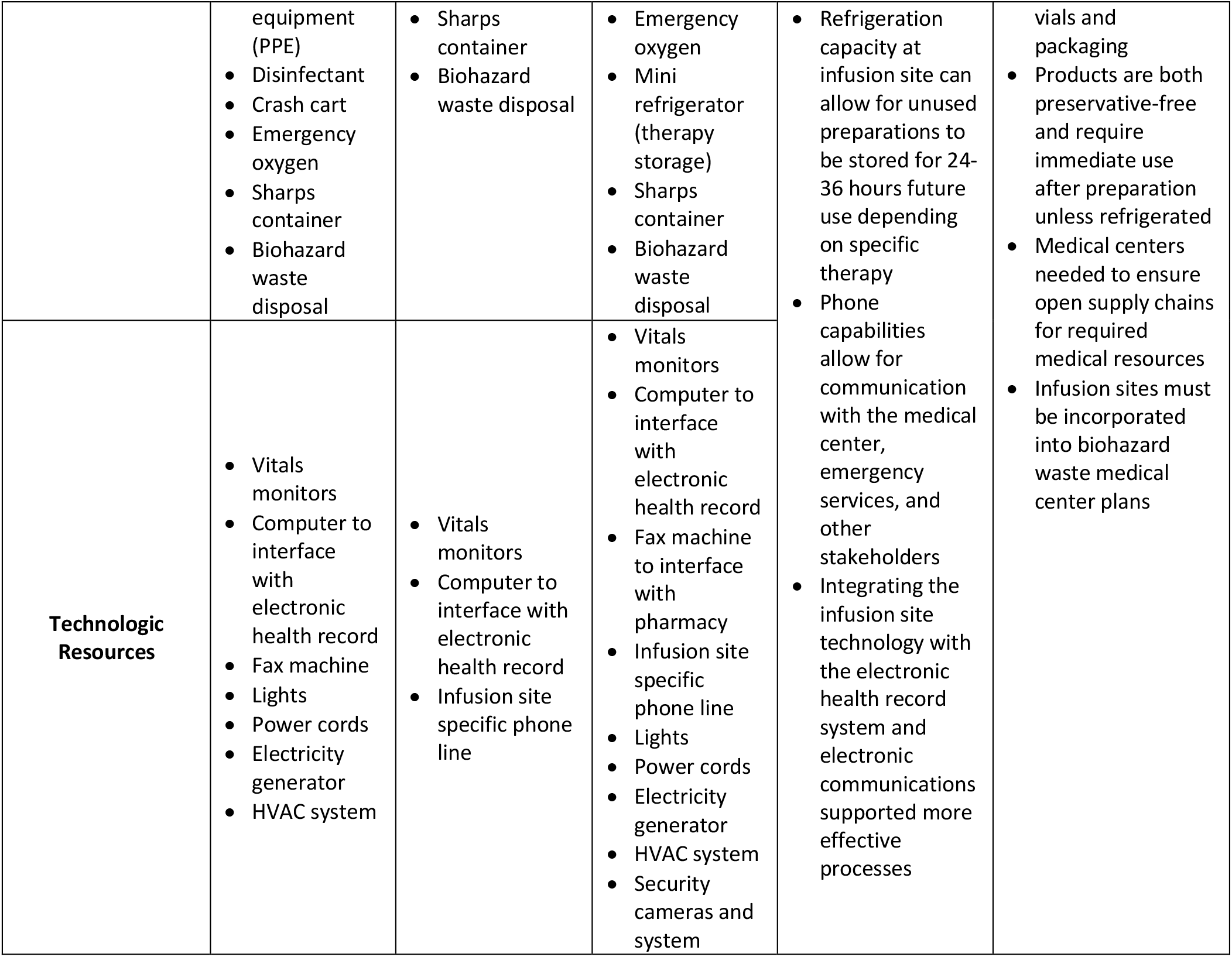
Monoclonal antibody infusion process physical environment and resource metrics from the three National Disaster Medical System-supported infusion sites and related strengths and challenges to inform implementation.

**Table 4.** Monoclonal antibody infusion process resilience, monitoring, and engagement metrics from the three National Disaster Medical System-supported infusion sites and related strengths and challenges to inform implementation.

| Resilience, Monitoring, & Engagement | Site 1 | Site 2 | Site 3 | Implementation Considerations |  |
| --- | --- | --- | --- | --- | --- |
|  |  |  |  | Strengths | Challenges |
| **Potential Adverse Events Protocol** | Crash-cart located within the tent<br>Site located adjacent to Emergency Department (ED) to address potential adverse events | Crash-cart located within the tent<br>Offsite of main medical campus, must call 911 for adverse events or related-emergencies | Crash-cart located within the tent<br>Site located adjacent to ED to address potential adverse events | Strong engagements with the local community members, providers, and other medical sites built trust and increased therapeutic demand<br>Utilizing an infusion dashboard and daily data metrics supported productive monitoring and evaluation<br>Infusion site proximity to ED optimized rapid care for adverse events<br>Dose repurposing or dose storage plan critical to address schedule and logistical disruptions<br>Infusion site processes integrated into the pre-existing medical center pandemic response ecosystem | Difficult to engage and build trust with particular patient and vulnerable communities due to mis- and disinformation on the COVID-19 pandemic<br>Pandemic strain and fatigue served as barriers to engaging providers<br>Barrier to stronger patient and community engagement was the delay in monoclonal antibody effectiveness data in outpatient populations |
| **Schedule Disruption Impacts** | Lacked pre-established schedule | Doses from scheduled patients who do not arrive were stored in refrigerator for next infusion appointment block within 24 hours | Doses from scheduled patients who do not arrive are stored in refrigerator for next infusion appointment block within 24 hours |  |  |
| **Monitoring & Evaluation of Infusion Site** | No formal monitoring and evaluation tools | Utilized dashboard and electronic health records to monitor and evaluate progress and adjust process | Uses whiteboard and electronic health records to monitor, evaluate, and adjust infusion process and schedule |  |  |
| **Patient Engagement** | Social media engagement such as Facebook Live<br>Local billboards and newspaper articles | Newspaper and online media<br>Provider referral system | Newspaper and online media<br>News media interviews<br>Provider referral system |  |  |
| **Provider Engagement** | Paper-based referral forms sent to provider offices | Provider and urgent care sites via email, fax, and phone | Provider and urgent care sites via email, fax, and phone |  |  |

### Resilience, Monitoring, and Engagement Metrics

Sustaining infusion sites through the pandemic required process resilience, monitoring, and engagement. Two major barriers that can affect process resilience were monoclonal antibody infusion-related adverse events and disruptions to the infusion schedule. The three sites had comprehensive plans and resources in place to address a potential adverse event including the presence of a crash cart at the infusion site, availability of oxygen, patient transport equipment, and medications to treat allergic reactions. The temporary tent sites were also placed adjacent to the ED of the medical centers to ensure close proximity to emergency services if needed. This was a challenge for the offsite physical environment of Site 2 as emergency services would need to be called in the event of an adverse reaction requiring further medical assistance. Disturbances to the schedule were not a potential challenge for Site 1 as it was walk-in based including referrals of ED patients. Sites 2 and 3 emphasized the importance of quickly refrigerating or relabeling an unused monoclonal antibody dose due to patients not arriving for their appointments. This proved to be a difficulty for sites on Fridays as they were closed on the weekends and the preservative-free monoclonal antibody drug products must be infused within 24 hours of preparation. Infusion process monitoring and evaluation varied greatly from site to site: one site did not conduct any real-time analysis and other sites implemented dashboards to monitor progress such as average patients per day, tracking adverse events, and patient appointment time estimates. A large barrier to monoclonal antibody infusion site implementation during the COVID-19 pandemic was engagement with patients and providers for education, outreach, and referrals.

## DISCUSSION

In these three Assistant Secretary for Preparedness and Response-supported monoclonal antibody infusion sites, our primary finding was that existing processes do not need to be reinvented to implement a successful infusion site during public health emergencies, as the therapy lends itself well to integration into existing outpatient infusion processes and ED/Urgent Care medical visits. The sites implemented various personnel, equipment, and resources to provide monoclonal antibody therapies in communities with large burdens of COVID-19. The general structures of the three monoclonal antibody process workflows described here are similar and have consistent major compartmental steps. Process variations were introduced to address state and local requirements on staffing, prescription orders, and to maintain medical center integration with other COVID-19 response workflows. As the COVID-19 pandemic and EUAs evolve, infusion site implementation and maintenance must remain adaptable to changes in therapeutic administration, clinical criteria, requirements, resources, and site needs.

Although a successful monoclonal antibody infusion site can be implemented with minimal staffing needs from the underlying healthcare system, the physical environment, resources, and work require planning and systems integration to ensure effectiveness, robust infection control, and safety. Medical volunteers or local paramedics can aid in staffing needs and also reduce the burden on the healthcare system during an emergency. The major strengths of these diverse sites derived from strong community and medical provider engagement on monoclonal antibodies, resilience to process disruptions, and optimized workflows of separating pre-infusion tasking and infusion-related activities between two nursing teams. The three sites demonstrated successful implementation during a pandemic through strong leadership and staff, collaboration with the National Disaster Medical System (NDMS), and flexibility to test and evaluate infusion process workflows. Common barriers and challenges across the sites included coordinating the preparation of the monoclonal antibody in the pharmacy, as it was not prepared at bedside. However, it is important to note that the EUA allows for the therapy to be prepared at bedside and this preparation mechanism may be more effective at particular types of sites, such as nursing homes, and at-home infusions. Infusion sites that scheduled patients were better able to address this barrier by batch preparing infusion bags and storing in a refrigerator. Scheduling monoclonal antibody infusion appointments was time- and staff-intensive; however, scheduling enabled more efficient workflows and monoclonal antibody preparation.

Confirming patient test positivity and scheduling individuals within 10 days of their symptom onset was another barrier to optimal monoclonal antibody infusions. Rigorous and timely testing and result communication was a necessary foundation for infusion site success due to the requirement for evidence of a positive test result. Future EUA changes and additional authorizations may address some of the logistical challenges and barriers in infusion site implementation such as reducing infusion times, changing storage and preparation requirements, and expanding patient criteria. Demand for this therapy has not yet been maximized in many communities and the sites’ process workflows can accommodate more patients than their average numbers. Community and provider engagement is critical for any new public health measure, but even more so during a pandemic, as the three sites reported challenges addressing misinformation and disinformation on COVID-19 treatments and control in their local communities.

The limitations of this descriptive analysis are rooted in its small sample size of three sites and limited geographic scope. However, this study has been uniquely conducted during the pandemic to inform ongoing public health action and infusion site implementation during this emergency. These therapies are not yet widely available internationally and lessons learned now in the U.S. may be generalizable to other settings implementing monoclonal antibodies for an emerging infectious disease.

## RECOMMENDATIONS FOR CURRENT AND FUTURE USE OF MONOCLONAL ANTIBODIES

The monoclonal antibody infusion site process description and assessment has informed general recommendations for the current implementation and future use of these therapies to tackle public health emergencies (Table 5). For current and future use, infusion process workflow and environment adaptability are critical as infusion times, requirements, and staffing change in emergencies. A primary recommendation is to build workflows that can be sustainably maintained in existing pandemic response ecosystems. Optimal staffing models require the minimal number of individuals with the appropriate targeted skills. Medical volunteers, paramedics, and other medical emergency support staff can be leveraged from local services to reduce the burden on the health system. In public health emergencies, it is important to innovatively expand potential monoclonal antibody administration sites beyond traditional settings.

**Table 5.** Monoclonal antibody infusion therapy and process recommendations for the COVID-19 pandemic and future emerging public health threats.

| Monoclonal Antibody Recommendation | Description |
| --- | --- |
| **Incorporate monoclonal antibodies into pandemic preparedness and response and existing health systems as an early intervention** | Monoclonal antibodies can:<br>• Be manufactured rapidly after neutralizing antibody identification<br>• Provide immediate immunologic support when other medical counter measures (MCMs) are under development or require time to achieve full effectiveness such as vaccines<br>• Serve as prophylaxis for individuals at high risk for infection<br>• Adapt to many forms of deployment during a public health emergency<br>• Integrate into existing health system processes such existing outpatient infusion processes and ED/Urgent Care med visits |
| **Strengthen process workflow and environment flexibility during public health emergency** | • Adjust monoclonal antibody administration process to policy changes<br>• Critical to monitor and evaluate process workflow to optimize and remain flexible to public health emergency conditions<br>• Adapt monoclonal antibody administration environment to infection control, weather, drug, and staffing changes |
| **Adapt staffing models to minimize burden, and maximize targeted skills** | • Establish workflow with minimal staffing needs<br>• Balance staffing needs with other emergency response activities<br>• Integrate non-traditional healthcare workers such as medical volunteers and paramedics |
| **Infusion site location expansion and innovative administration** | • **Community-based sites:** multiple medical centers partner to implement a monoclonal antibody infusion site, share resources and staffing, and minimize individual burden<br>• **Rapid testing adjacent sites:** co-locate monoclonal antibody site with rapid testing capabilities to refer and immediately treat patients<br>• **Car-based infusion or injection:** alleviate the physical environment by delivering monoclonal antibodies and observing patients in cars<br>• **Home administration:** administer monoclonal antibodies in patients' homes<br>• **Nursing homes:** administer monoclonal antibodies in nursing homes or long-term care facilities |
| **Ensure strong engagement and equity** | • Engage with local communities to dispel mis- and disinformation regarding treatments<br>• Empower communities and providers with the knowledge of new therapeutic options and impact data<br>• Ensure monoclonal antibody allocation equity by directing information to populations that are vulnerable, most in need, and likely to meet eligibility criteria |
| **Improved therapy formulations and delivery mechanisms** | • Expand routes of drug administration (e.g., intramuscular, subcutaneous)<br>• Minimize temperature stability and drug product preparation requirements |
| **Standard data collection and effectiveness study integration for outpatients** | • Establish data collection standards for early adopters of monoclonal antibody infusion to permit rapid assessment and large-scale evaluation<br>• Pair monoclonal antibody distribution with data collection network to better understand the therapeutic impact during EUA periods |
| **Sustainable use and public health integration through other disease targets** | • Promote monoclonal antibodies in emerging infectious disease preparedness and response toolkit<br>• Build upon the therapeutics momentum from the pandemic<br>• Continue innovative monoclonal antibody research and study delivery mechanisms and emergency implementation techniques<br>• Partner with organizations researching the application of monoclonal antibodies for other disease targets and public health threats |

A future outbreak or pandemic could be ignited by a more transmissible pathogen, in which it would be prudent to further minimize staff and patient interactions. One potential solution is patient infusion or injection of a monoclonal antibody therapy with observation in patients’ vehicles, decreasing interactions in a physical environment, space, and indoor infection control systems. This intervention may not be suitable for all settings and vulnerable populations, but it can reduce the strain on physical environments and decrease potential transmission events between patients and health care workers. Further integration of monoclonal antibody delivery into communities could occur by co-locating infusion sites with rapid testing sites so that patients notified of positivity and meeting eligibility criteria could easily access treatment. Infusions and injections may also be administered in the home, removing the need for a physical environment, but potentially increasing the staffing needs and time. As novel treatments arise, such as monoclonal antibodies, strong engagement with the public and equitable distribution of such therapeutics to vulnerable populations is critical.^14^ Currently, monoclonal antibodies are delivered via intravenous infusion; however, research may soon enable intramuscular and subcutaneous delivery.^15^ There is evidence that current monoclonal antibody therapies may show reduced neutralization and potential effectiveness against novel SARS-CoV-2 virus variants to which the drugs were not optimized.^16^ However, a strength of monoclonal antibodies is rooted in their adaptability and rapid production. Monoclonal antibody therapies can act as a platform biologic that can be updated as emerging infectious diseases evolve and evade targeting.

Measuring the effectiveness of new therapies, especially in outpatient populations, during a public health emergency is difficult because resources are focused on saving lives. Establishing site data collection standards to rapidly assess effectiveness and pairing this with the early distribution of new therapies during an emergency, such as monoclonal antibodies, would improve large-scale evaluation. Implementation lessons learned can be translated for the next pandemic. Innovative research, delivery mechanisms, and implementation techniques for monoclonal antibodies must be further studied and optimized, and this can be accomplished through the lens of other pathogens and public health threats. The emerging infectious disease preparedness and response toolkit is growing to incorporate monoclonal antibodies and building upon the therapeutics momentum in the current pandemic is important for the next pandemic.

## Supporting information

SQUIRE 2.0 Checklist

## Data Availability

The data was collected as part of the ASPR public health response to the COVID-19 pandemic and is not publicly available.

## ACKNOWLEDGEMENTS

The authors acknowledge the significant efforts of the members of the Disaster Medical Assistance Teams (DMAT) who coordinated the monoclonal antibody infusion site set-up, initiation, and integration with the collaborating medical centers. The authors would also like to thank the incredible infusion site leadership, implementation, and clinical staff at El Centro Regional Medical Center (El Centro, CA), TMC HealthCare (Tucson, AZ), and Sunrise Hospital and Medical Center (Las Vegas, NV), who provided extensive time and support for this study.

