## Supplementary Information for "Implementation of SARS-CoV-2 monoclonal antibody infusion sites at three medical centers in the United States: Strengths and challenges assessment to inform COVID-19 pandemic and future public health emergency use"

**TITLE**

Anastasia S. Lambrou, PhD

### SUPPLEMENTARY INFORMATION

**SI Table 1.** Monoclonal antibody US Food and Drug Administration (FDA) Emergency Use Agreement EUA prospective patient criteria as of February 2021.

| Prospective Patient Age | Risk Factors |
|---|---|
| **≥18 years of age** *and at least one of the following risk factors* | • Have a body mass index (BMI) ≥35<br>• Have chronic kidney disease<br>• Have diabetes<br>• Have immunosuppressive disease Are currently receiving immunosuppressive treatment<br>• Are ≥65 years of age |
| **≥55 years of age** *and at least one of the following risk factors* | • Cardiovascular disease<br>• Hypertension<br>• Chronic obstructive pulmonary disease/other chronic respiratory disease |
| **12 – 17 years of age** *and at least one of the following risk factors* | • BMI ≥85th percentile for their age and gender based on CDC growth charts<br>• Sickle cell disease<br>• Congenital or acquired heart disease<br>• Neurodevelopmental disorders, for example, cerebral palsy<br>• Medical-related technological dependence, for example, tracheostomy, gastrostomy, or positive pressure ventilation (not related to COVID-19)<br>• Asthma, reactive airway or other chronic respiratory disease that requires daily medication for control |

**SI Table 2.** Monoclonal antibody infusion site process assessment metric descriptions and corresponding semi-structured interview questions.

| Metric | Definition | Data |
|---|---|---|
| **Engagement & Workflow Logistics** | Components, steps, and official procedures of engaging with all stakeholders and the workflow of the infusion process to assess areas of improvement. | List the steps of the infusion process from recruitment to scheduling and receiving the therapy |
| | | How long does each step take from the perspective of patients and staff? |
| | | What are the current patient/community engagement/outreach mechanisms? |
| | | What are the current provider/clinical engagement/outreach mechanisms? |
| | | What forms of transportation do patients use to travel to and from their appointment? |
| | | What barriers do patient face when scheduling or attending the appointment? |
| | | How can the site increase provider/prescriber buy-in? |
| | | What are the hard vs. soft constrains of the process? |
| **Timing** | Metrics related to timing of steps and engagement and therapy-quality related metrics such as preparation and temperature. | Time from registration/scheduling to appointment (mean, individual data if available) |
| | | Time from symptom onset to infusion |
| | | Time from positive test to infusion |
| | | Total appointment time |
| | | Estimated time for each infusion process step |
| | | Transition time between patients/infusions |
| | | Infusion site hours of operation |
| | | Infusion site staff shift hours/timing |
| | | List mAb therapy quality assurance process |
| | | Pharmacist preparation timing and location |
| | | Average amount of time between pharmacist preparation/cooling conditions to infusion |
| **Staffing** | Information on staff, their roles, and requirements to inform staffing models and process improvements. | List the number and types of workers maintaining the infusion site |
| | | What is the level of training needed for specific infusion site workers? |
| | | Is there a minimum staffing requirement? |
| | | Is there a maximum number of individuals allowed in the space at once? |
| | | What is the role of each type of worker? |
| | | Can any worker take on multiple roles? |
| | | How does process timing and steps change if the staff is down one nurse/physician/etc.? |
| | | Which process steps are staff dependent? |
| | | What is the most labor-intensive step for workers? |
| | | Which process step requires the most skills/training? |
| **Physical Environment** | Information, layout, and interactions within the facilities/physical | Map infusion site and logistics environment |
| | | Locations/direction of chairs/patients |
| | | Dimensions of infusion site |

| | | |
|---|---|---|
| | environment in the infusion site to inform process improvements and prototype set-ups. | Infection control/distancing in physical environment |
| | | Can the size or shape of the space be altered? |
| | | Is any daily setup or deconstruction required? |
| **Resources** | All resources needed that are not staff to inform process model, scale-up, and process improvements. | List all physical resources needed for the infusion process |
| | | PPE resources needed |
| | | Infusion site/tent resources |
| | | Information systems resources |
| | | Could the patient be observed in another setting, i.e., car? What parts of the process do not have to occur in the tent/site? |
| **Monitoring & Resilience** | Flexibility and resilience of system to process disturbances, emergencies, and capacity changes. | What happens if an emergency/adverse event occurs? |
| | | How common are emergency/adverse events and how do they impact staffing/flow/etc.? |
| | | What happens if a patient does not show up for their appointment? |
| | | How common are "no shows" and how do they impact staffing/flow/etc.? |
| | | Describe the system that is currently being used for patient scheduling |
| | | Describe the system currently used to monitor and evaluate (M&E) the infusion process |
| | | Describe the system that is currently being used to |
| | | Describe the system that is currently being used to monitor logistics and resource allocation |

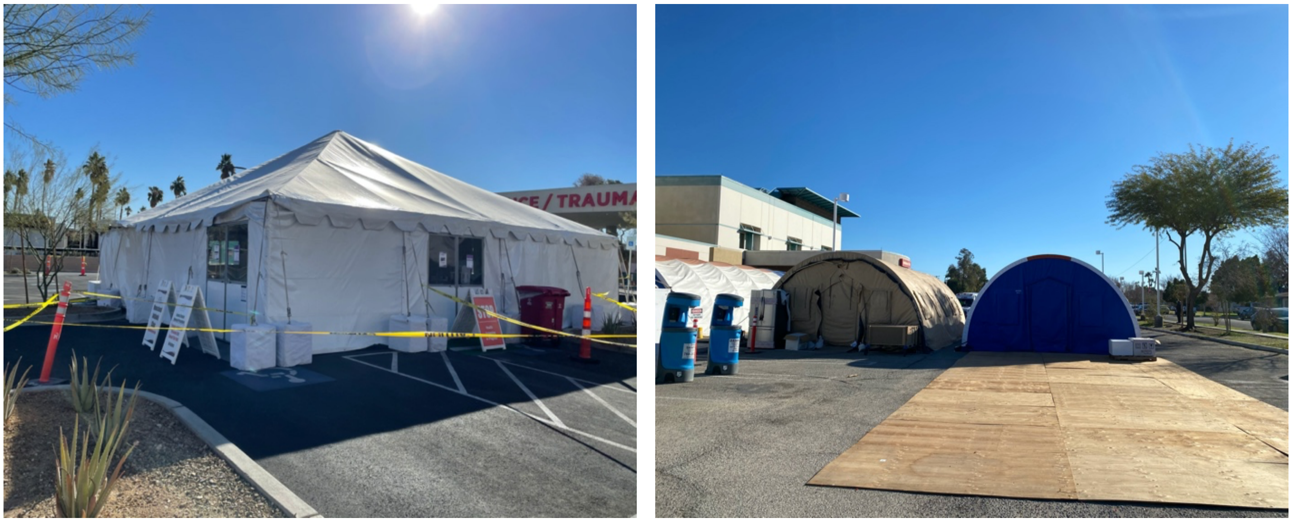

**SI Figure 1.** External physical environments of the temporary tent monoclonal antibody infusion sites.

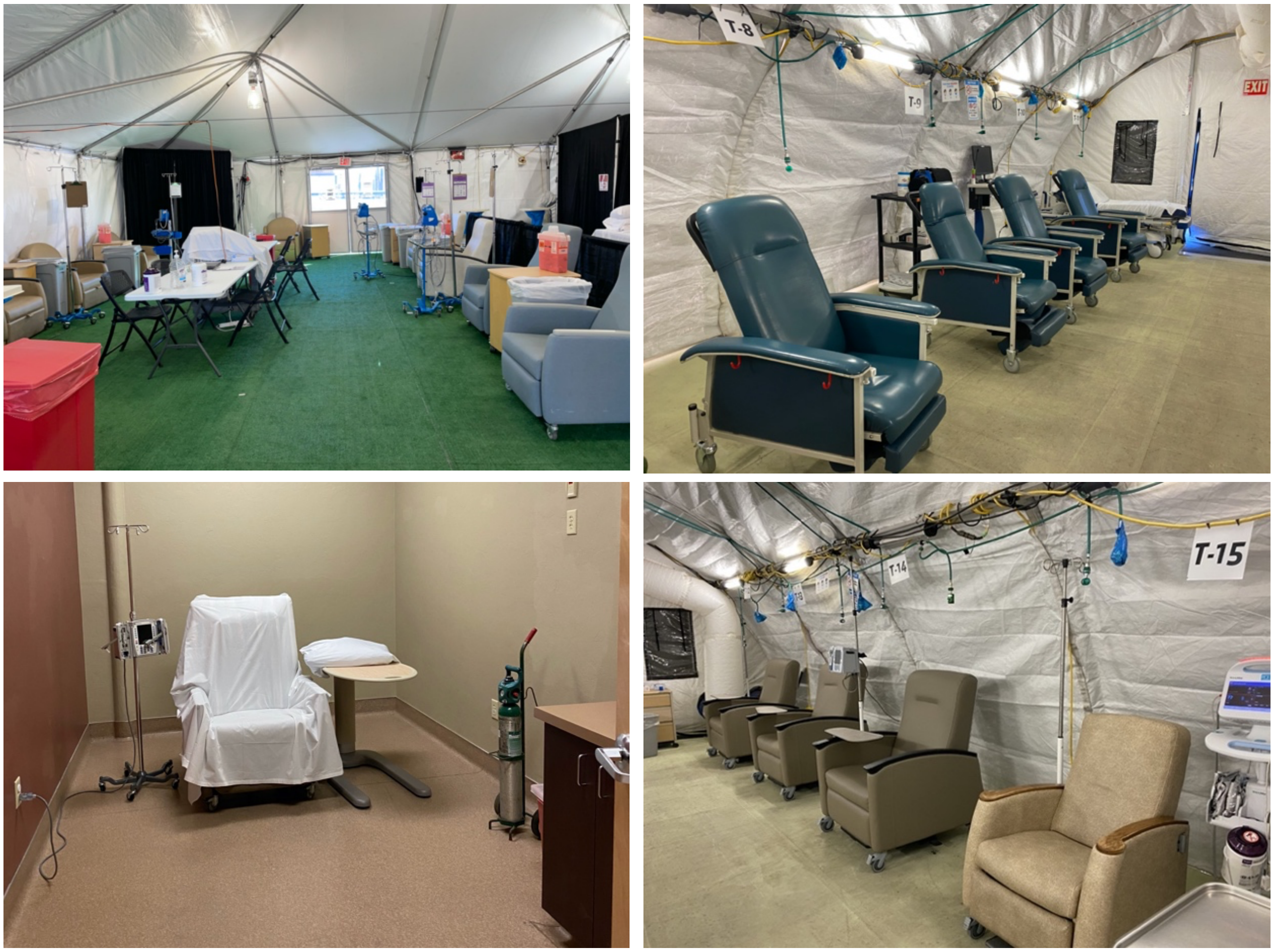


**SI Figure 2.** Examples of the internal physical environments of the three monoclonal antibody infusion sites exhibiting both open-concept and individual patient room layouts for the infusion process
