## Supplementary material for "Implementation of SARS-CoV-2 monoclonal antibody infusion sites at three medical centers in the United States: Strengths and challenges assessment to inform COVID-19 pandemic and future public health emergency use": SQUIRE 2.0 Checklist

### Revised Standards for Quality Improvement Reporting Excellence (SQUIRE 2.0) Author Checklist

| Text Section and Item Name | | Corresponding Manuscript Page Number |
|---|---|---|
| **Title and Abstract** | | |
| 1 | Title | 1 |
| 2 | Abstract | 2 |
| **Introduction** | | |
| 3 | Problem Description | 4-5 |
| 4 | Available Knowledge | 4-5 |
| 5 | Rationale | 5 |
| 6 | Specific aims | 5 |
| **Methods** | | |
| 7 | Context | 6 |
| 8 | Interventions | 6-7 |
| 9 | Study of Interventions | 6-7 |
| 10 | Measures | 7 |
| 11 | Analysis | 8 |
| 12 | Ethical Considerations | 6 |
| **Results** | | |
| 13 | Results | 8-19 |
| **Discussion** | | |
| 14 | Summary | 20 |
| 15 | Interpretation | 20-21 |
| 16 | Limitations | 21 |
| 17 | Conclusions | 22-24 |
| **Other Information** | | |
| 18 | Funding | 25 |
